# Efficacy of vaccination against severe COVID-19 in relation to Delta variant and time since second dose: the REACT-SCOT case-control study

**DOI:** 10.1101/2021.09.12.21263448

**Authors:** Paul M McKeigue, David A McAllister, Sharon J Hutchinson, Chris Robertson, Diane Stockton, Helen M Colhoun, for the PHS COVID-19 Epidemiology and Research Cell

**Affiliations:** Usher Institute, College of Medicine and Veterinary Medicine, University of Edinburgh, Teviot Place, Edinburgh EH8 9AG, Scotland; Institute of Health and Wellbeing, University of Glasgow, 1 Lilybank Gardens, Glasgow G12 8RZ; School of Health and Life Sciences, Glasgow Caledonian University. SH - Professor of Epidemiology and Population Health; Department of Mathematics and Statistics, University of Strathclyde, 16 Richmond Street, Glasgow G1 1XQ; Institute of Genetics and Cancer, College of Medicine and Veterinary Medicine, University of Edinburgh, Western General Hospital Campus, Crewe Road, Edinburgh EH4 2XUC, Scotland; Public Health Scotland, Meridian Court, 5 Cadogan Street, Glasgow G2 6QE

## Abstract

**Background:** The objectives of this study were to investigate whether vaccine efficacy against severe COVID-19 has decreased since Delta became the predominant variant; (2) whether efficacy wanes with time since second dose.

**Methods:** Efficacy was estimated in a matched case-control study that includes all diagnosed cases of COVID-19 in Scotland up to 19 August 2021. The main outcome measure was severe COVID-19, defined as cases with entry to critical care or fatal outcome.

**Findings:** Efficacy of vaccination against severe COVID-19 decreased in May 2021 coinciding with the replacement of the B.1.1.7 (Alpha) by the B.1.617.2 (Delta) variant in Scotland, but this decrease was reversed over the next month. In the most recent time window, the efficacy of two doses was 91% (95 percent CI 87% to 94%) for the AstraZeneca product and 92% (95 percent CI 88% to 95%) for mRNA (Pfizer or Moderna) products. Efficacy of the AstraZeneca product against severe COVID-19 declined with time since second dose to 69% (95 percent CI 52% to 80%) at 20 weeks from second dose. Efficacy of mRNA vaccines declined in the first ten weeks from second dose but more slowly thereafter to 93% (95 percent CI 88% to 96%) at 20 weeks from second dose.

**Interpretation:** These results are reassuring with respect to concerns that efficacy against severe COVID-19 might have fallen since the Delta variant became predominant, or that efficacy of mRNA vaccines wanes with increasing time since second dose. However it is now clear that efficacy of the AstraZeneca vaccine against severe COVID-19 wanes substantially by 20 weeks from second dose, suggesting that delivery of booster doses should initially focus on those who received this type of vaccine.

**Funding:** No specific funding was received for this work. HC is supported by an endowed chair from the AXA Research Foundation.

**Research in context:** *Evidence before this study:* Several reports have suggested that efficacy of COVID-19 vaccines has fallen since the Delta variant became predominant, or that efficacy wanes with time since second dose. The starting point for this study was the evidence of waning efficacy cited by the CDC, the FDA and more recently the UK Joint Committee on Vaccination and Immunisation in support of their recent recommendations for delivery of booster doses for the general population.

*Added value of this study:* This study shows that efficacy of both AstraZeneca and mRNA vaccines against severe COVID-19 (fatal or requiring critical care) remains high (around 90%) in the most recent time window, but that efficacy of the AstraZeneca vaccine wanes to about 70% by 20 weeks from second dose. In contrast efficacy of the mRNA vaccines wanes rapidly at first but stabilises at about 90% by 20 weeks from second dose.

*Implications of all the available evidence:* These results suggest that booster doses of vaccine are not needed for those who have received two doses of mRNA vaccine, except for vulnerable individuals who may require a third primary dose.

## Introduction

Recent reports have suggested that efficacy of vaccines against COVID-19 infection may have fallen since the B.1.617.2 (Delta) variant became predominant [1–6]. Other studies have raised concerns that efficacy may wane with time since second dose [3,7,8]. These concerns have led US and UK advisory bodies [9,10] to recommend booster doses for the general population.

Studies of efficacy against infection are subject to ascertainment bias unless they are based on testing at predetermined regular intervals [3]. Studies of efficacy against severe COVID-19, defined as cases that are fatal or require critical care, are less susceptible to ascertainment bias and this is also the outcome most relevant to health-care capacity. The objectives of this study were to investigate for the two main classes of vaccine: (1) whether efficacy against severe COVID-19 has decreased since Delta became the predominant variant; (2) whether efficacy of two doses against severe COVID-19 wanes with time since second dose.

## Methods

The design of the REACT-SCOT case-control study has been described in detail previously [11–13]. In brief, for every incident case of COVID-19 in the Scottish population ten controls not previously diagnosed with COVID-19, matched for one-year age, sex and primary care practice and alive on the day of presentation of the case that they were matched to were selected using the Community Health Index database. Diagnosed cases of COVID-19 were those with a positive nucleic acid test, a hospital discharge diagnosis coded as COVID-19, or a death certificate with mention of COVID-19. As previously, to minimise ascertainment bias we pre-specified the primary outcome measure as severe COVID-19, defined as diagnosed cases with entry to critical care within 21 days of first positive test, death within 28 days of first positive test or any death for which COVID-19 was coded as underlying cause [11]. We also examined as a secondary outcome the broader category of hospitalised or fatal COVID-19, with hospitalisation defined as admission within 14 days of first positive test. Though the data extract included cases presenting up to 2 September 2021, the analyses reported here are restricted to cases and controls presenting from 1 December 2020 to 19 August 2021, ensuring follow-up for at least 14 days after presentation date to allow classification of hospitalisation and (for most cases) severity based on entry to critical care or fatal outcome.

The vaccination programme in Scotland began on 8 December 2020. By 24 March 2021 half the adult population had received a first dose, and by 7 June half the adult population had received a second dose [14]. Vaccination status was defined by the number of doses received at least 14 days before presentation date.

The incidence density sampling design controls not only for the matching factors of age, sex and primary care practice but also for calendar time. Rate ratios for severe COVID-19 were estimated from conditional logistic regression models. The efficacy of vaccination is 1 minus the rate ratio. Covariates included in each model were those previously identified as strong predictors of severe disease in this population: care home residence, risk category (no risk condition, moderate risk condition, clinically extremely vulnerable), number of non-cardiovascular drug classes dispensed in last 240 days and recent hospital stay [11–13]. The criteria used to classify individuals as having conditions designated as moderate risk [11] or as clinically extremely vulnerable (and thus eligible for shielding) [13] have been described in detail previously.

To investigate the effect of the Delta variant we examined how efficacy varied with calendar time, and to investigate possible waning we examined how efficacy varied with time since last dose. To show these relationships without predefined categories, the initial analysis presents line plots of log rate ratios estimated within sliding 42-day time windows against calendar time and against time since last dose. These sliding time windows were used only to generate these plots. For a formal comparison between time periods before and after Delta became the predominant variant, we estimated rate ratios before and after 19 May 2021: the date on which Delta became the main variant in Scotland [15].

A key question for policy is whether the early waning of vaccine efficacy after the second dose tapers off after a few months. To investigate this we compared the fit of two families of model: (1) a “waning to zero efficacy” model in which the effect of vaccination on the scale of log rate ratio decays exponentially to zero with time since second dose; (2) a “waning to constant efficacy” model in which the effect of vaccination is the sum of two terms: a time-invariant effect and a waning effect that decays exponentially with time since second dose. For each of these two model families, a model was fitted for each value of the decay half-life over a sequence of values from 10 to 500 days and a profile likelihood confidence interval for the half-life was obtained as the range of half-life values over which the log-likelihood of the model was within 1.92 natural log units of its maximum value. Comparison between the best-fitting waning to zero model and the best-fitting “waning to constant efficacy” model was based on the difference in log-likelihood between these nested models.

## Results

Tables S2 and S3 show the distributions of risk factors in cases and their matched controls, for the 5644 severe cases and the 21671 cases in the broader category of hospitalised or fatal. These results are provided for reference only: the reader is cautioned that unconditional odds ratios calculated from these tables cannot be used to estimate rate ratios because of the matched design [16,17]. Over all cases and matched controls, the median and interquartile range of the time since last dose was 40 (24 to 62) days for those who had received a single dose and 97 (62 to 132) days for those who had received two doses. Of those who had received two doses of an mRNA vaccine by the date of the latest extract, 9% had received the Moderna product.

### Vaccine efficacy before and after the Delta variant became predominant

Figure 1 (a) shows that the rate ratio for severe COVID-19 associated with a single dose of vaccine in June to July 2021, after the Alpha variant was replaced by the Delta variant, was similar to that in March to April. There is however an obvious blip, with a temporary increase in the rate ratio (corresponding to a decline in efficacy) from early May to early June. The rate ratios for severe COVID-19 associated with two doses of vaccine show a similar perturbation during May 2021, but the estimates of rate ratios for time windows before May 2021 are imprecise because at this time few individuals had received their second dose. To compare the rate ratio before and after the date that the Delta variant became predominant, a conditional logistic regression model was fitted with the effect of two doses versus none nested within each level of an indicator variable defined as presentation date on or after 19 May 2021. The rate ratio for severe disease associated with two doses of vaccine was 0.05 (95% CI 0.01 to 0.27, *p*=4 × 10^*-*4^) before 19 May and 0.09 (95% CI 0.07 to 0.11, *p*=7 × 10^*-*125^) from 19 May onwards. The confidence interval for the rate ratio before 19 May is wide because few individuals had received two doses before April 2021 and there were few severe cases from April to mid-May 2021. Figure 1 (b) shows that the rate ratio for the broader category of hospitalised or fatal disease associated with two doses of vaccine increased (and thus efficacy was lower) after the Delta variant became predominant. The rate ratio was estimated as 0.11 (95% CI 0.07 to 0.19, *p*=8 × 10^*-*17^) before 19 May and 0.17 (95% CI 0.16 to 0.18, *p*=1 × 10^*-*404^) from 19 May onwards

**Fig 1.**
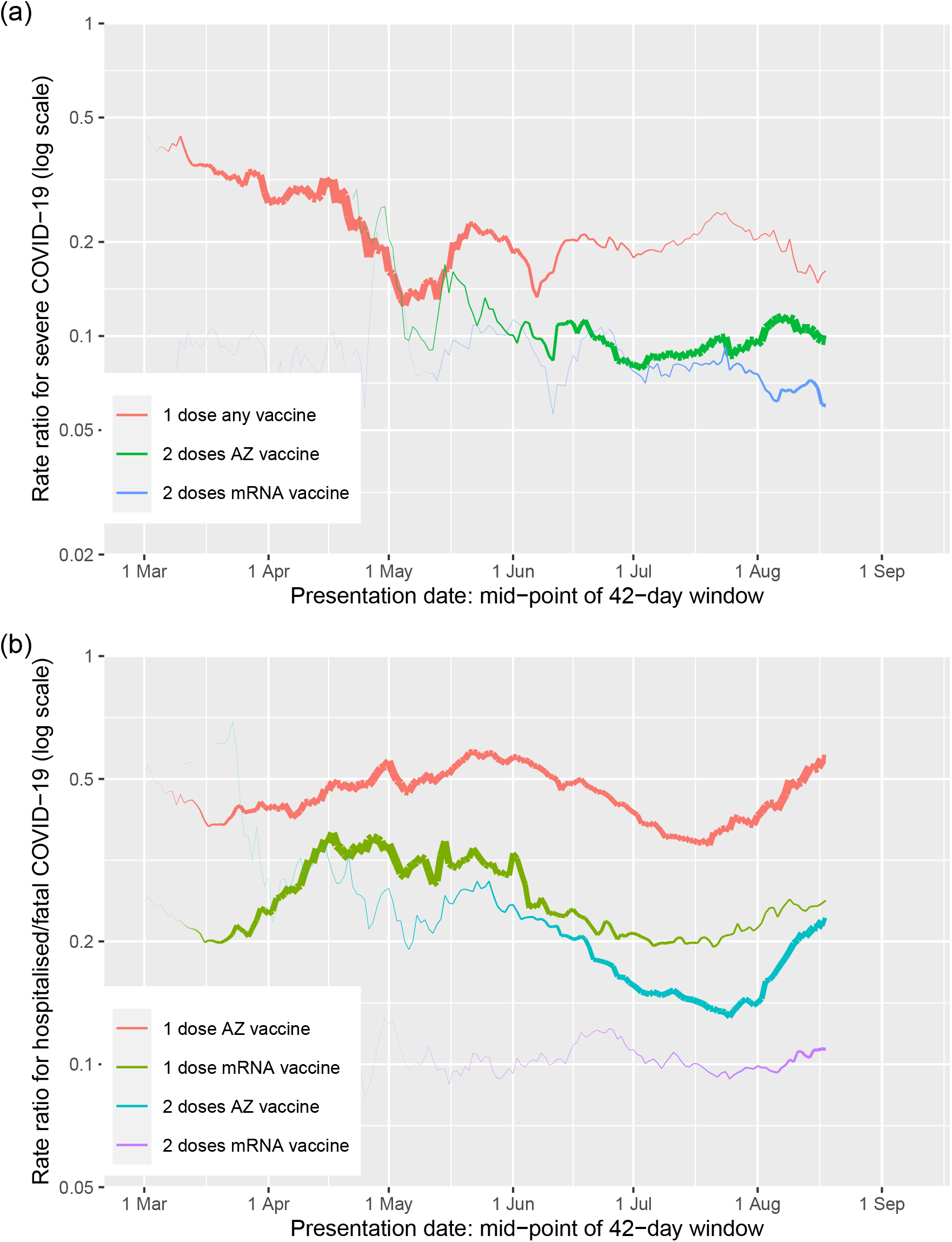
Relation of vaccine efficacy to calendar time: (a) severe COVID-19 (single-dose categories for AZ and mRNA vaccines have been combined as the numbers of events in those with 1 dose of mRNA vaccine were small); (b) hospitalised or fatal COVID-19. Rate ratios in conditional logistic regression model, adjusted for covariates. For each effect, line thickness is proportional to precision of estimate.

Figure 1 (a) shows that against severe COVID-19, the efficacy of two doses of the AstraZeneca and mRNA vaccines did not differ after May 2021, but Figure 1 (b) shows that against the broader category of hospitalised or fatal COVID-19 the AstraZeneca vaccine had lower efficacy than the mRNA vaccines. In the most recent 42-day time window centred on 29 July 2021, the efficacy of two doses against severe COVID-19 was 91% (95 percent CI 87% to 94%) for the AstraZeneca product and 92% (95 percent CI 88% to 95%) for mRNA (Pfizer or Moderna) products. Against the broader category of hospitalised or fatal COVID-19, efficacy in this time window was slightly lower for the AstraZeneca product [86% (95 percent CI 83% to 88%)] than for mRNA vaccines [90% (95 percent CI 88% to 92%)].

### Vaccine efficacy by time since second dose

Figure 2 (a) shows that the log rate ratio for severe COVID-19 increases (and thus efficacy decreases) over the first ten weeks after the second dose for both AstraZeneca and mRNA vaccines. After this the slope of this relationship flattens for mRNA vaccines but not for the AstraZeneca vaccine. Figure 2 (b) shows the same analysis for the broader category of hospitalized or fatal COVID-19. In the 42-day time window centred on 20 weeks from second dose, the efficacy of the AstraZeneca product against severe COVID-19 is 69% (95 percent CI 52% to 80%) but the efficacy of mRNA vaccines is 93% (95 percent CI 88% to 96%). For efficacy against hospitalised or fatal COVID-19 at 20 weeks from second dose the corresponding estimates were 58% (95 percent CI 50% to 65%) and 89% (95 percent CI 86% to 91%).

**Fig 2.**
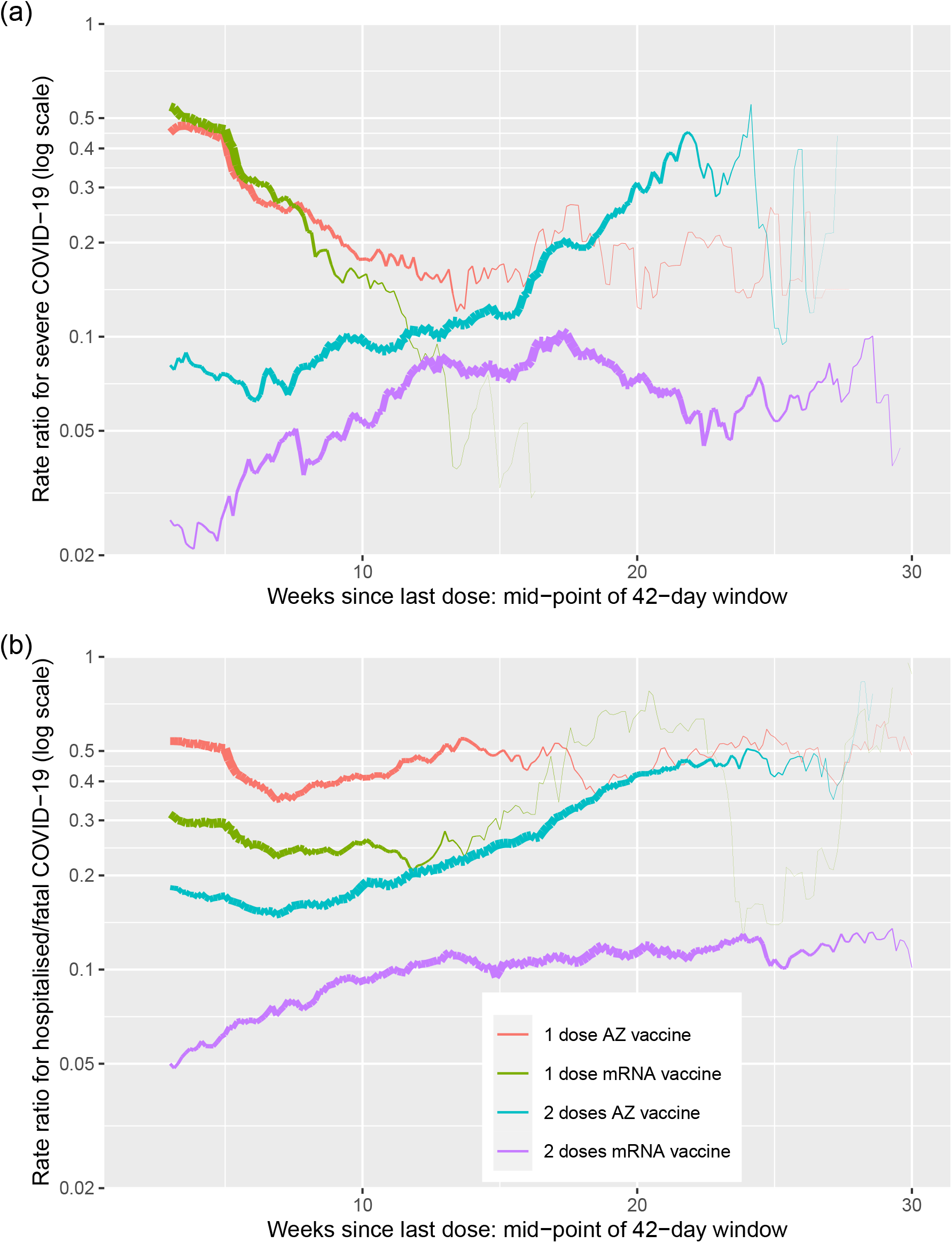
Relation of vaccine efficacy to time since last dose: (a) severe COVID-19; (b) hospitalised or fatal COVID-19. Rate ratios in conditional logistic regression model, adjusted for covariates. For each effect, line thickness is proportional to precision of estimate.

Figure S1 shows the relation of efficacy against hospitalisation to time since second dose by clinical risk category. The differences between the AstraZeneca vaccine and the mRNA vaccines in pattern of waning is evident in each risk category: thus these differences cannot be explained by the different risk profiles of those who received mRNA vaccines and those who received AstraZeneca vaccines.

Modelling of the relation of efficacy to time since second dose was based on comparison of “waning to zero efficacy” and “waning to constant efficacy” models as described above. Results are shown in Table S1. The decay curves corresponding to the best-fitting waning to constant efficacy model for each outcome are shown in Figure S2. For severe COVID-19, there was no clear evidence (difference in log-likelihood < 2) favouring waning to constant efficacy over waning to zero. For the broader category of hospitalised or fatal COVID-19, there was strong evidence favouring waning to constant efficacy over waning to zero. For mRNA vaccines the model best supported by the data was one in which efficacy was the sum of a rapidly waning effect with half-life of 16 (95% CI 4 to 101) days and a time-invariant efficacy of 93%. For the AstraZeneca vaccine the model with waning to constant efficacy was supported, but with a much longer decay half-life (lower bound 211 days) such that the decay curve up to 30 weeks is barely distinguishable from a straight line.

## Conclusions

### Statement of principal findings

- The efficacy of two vaccine doses against severe COVID-19 in the most recent time windows is around 92% and differs only slightly between AstraZeneca and mRNA products. Efficacy against the broader category of hospitalised or fatal COVID-19 is lower for the AstraZeneca vaccine than for mRNA vaccines (86% versus 90%). The replacement of the Alpha by the Delta variant in May 2021 was accompanied by a temporary reduction of efficacy against severe disease: a possible explanation is that the early spread of the new variant was associated with higher infecting doses as suggested by the lower PCR cycle threshold values recorded in surveillance data for England [18].
- Efficacy of mRNA vaccines against severe or hospitalised COVID declines during the first ten weeks after the second dose but stabilises thereafter at about 90%. Efficacy of the Astra Zeneca vaccine continues to decline to about 60% at 20 weeks. Although the log rate ratio is the natural scale on which to model these effects, we emphasize that changes on this scale do not equate to changes in absolute risk: thus a twofold increase in the rate ratio can represent a decline of efficacy from 97.5% to 95%, or a decline from 80% to 60%. It is natural to model waning as an exponential decay of the log rate ratio (a linear effect would imply that efficacy eventually becomes negative). Although a model in which efficacy against hospitalisation is the sum (on a scale of log rate ratio) of a rapidly-waning effect and a time-invariant effect is supported by these data, the underlying mechanism of this is not clear, as the rate of decline of neutralising antibodies induced by mRNA vaccines [19] appears too slow to explain the rapid decline of efficacy in the first two months since second dose.

### Strengths and limitations

Strengths of this study are the focus on severe COVID-19 as an outcome measure for which ascertainment is complete and ascertainment bias should be minimal, the elimination of confounding by calendar time in the matched case-control design, and the ability to control for confounding by comorbidities and recent inpatient stay through linkage to electronic health records. The incidence density case-control design excludes those who have previously tested positive for COVID-19: a study of reinfections will be reported elsewhere. Our estimates for waning of mRNA vaccines are based almost entirely on the Pfizer vaccine: only recently has the Moderna vaccine been used in Scotland.

For the secondary outcome of hospitalised or fatal COVID-19 the numbers of events are larger but we cannot easily distinguish admissions caused by COVID-19 from admissions for other conditions where a positive COVID test is an incidental finding on admission. Where hospitalisations with COVID-19 are misclassified as hospitalisations caused by COVID-19, efficacy of a vaccine against hospitalisation caused by COVID-19 may be underestimated if its efficacy against test-positive infection is lower than its efficacy against disease.

For investigating the possible effect of the Delta variant on efficacy, a limitation is that we do not have direct measurements of variant type; however in Scotland the Alpha variant was almost completely replaced by Delta over a few weeks in May 2021 [15], and the effect of this is visible in the time window plots as a temporary perturbation of efficacy. As few people had received their second dose before April 2021 and from April to May 2021 the number of severe cases was low, estimates of efficacy of two doses against severe COVID-19 are based mainly on cases occurring after May 2021. Although the effects of calendar time and time since second dose are confounded with other factors not considered in this analysis including seasonality, the build-up of natural immunity, and the changing morbidity profile of cases, the objective of this study is to establish whether efficacy is waning in the population as a whole and thus to lay an evidence base for policy.

### Comparison with other studies

Several studies have suggested that vaccine efficacy against infection may have fallen since Delta became the predominant variant, or that efficacy wanes with increasing time since second dose. For efficacy against infection, the most reliable evidence is from the UK Office of National Statistics (ONS) Covid-19 Infection Survey, based on regular monthly PCR testing: a study based on this reported that efficacy against infection had fallen from 79% to 67% for the AstraZeneca vaccine since Delta became the predominant variant, but had remained around 80% for the Pfizer vaccine [3]. Three other recent studies from the UK [2,15,20] using test-negative controls have estimated recent efficacy against symptomatic infection to be lower for the AstraZeneca vaccine than for the Pfizer vaccine.

Results of a recent study from the Kaiser Permanente Health Program showing that efficacy of the Pfizer vaccine against hospitalisation with COVID-19 remained around 90% after six months since second dose are consistent with our findings [21]. Few studies have compared the waning of efficacy of mRNA and AstraZeneca vaccines against hospitalisation. A recent study from Public Health England based on data up to 3 September 2021 estimated that efficacy against hospitalisation beyond 20 weeks from second dose remained around 92% for the Pfizer vaccine, but declined to 77% for the AstraZeneca vaccine [20]. Limitations of that study are the restriction to cases ascertained from community testing (Pillar 2) and the test-negative control design. In contrast we estimate that efficacy of the AstraZeneca vaccine against hospitalisation wanes to around 60% by 20 weeks.

### Policy implications

On the basis of reports that vaccine efficacy against SARS-CoV-2 had fallen since Delta became the predominant variant, and that efficacy waned with time since second dose, the CDC and FDA recommended booster doses for all adults in the US [9] though the Vaccines and Related Biological Products Advisory Committee subsequently limited their recommendation to those aged over 65 years. In the UK the JCVI recommended booster doses for all those aged over 50 years [10]. Others have expressed doubts about the evidence of waning efficacy, and argued that “currently available evidence does not show the need for widespread use of booster vaccination in populations that have received an effective primary vaccination regimen.” [22]. A report from Public Health England suggested that declining efficacy against infection might even be beneficial in the long term by “boosting the primed immune system of vaccinees who would experience mild or asymptomatic infections.” [23]

With respect to the Delta variant, our results are more reassuring than earlier reports, in that we find that while vaccine efficacy against severe COVID-19 declined when the Alpha variant was replaced by the Delta variant, this decline was only temporary. With respect waning with time since second dose, it is now clear that efficacy of the AstraZeneca vaccine against severe COVID-19 and hospitalisation wanes up to 20 weeks from second dose to about 60%, but efficacy of the mRNA vaccines appears to stabilise at about 90% after rapid waning in the first 10 weeks from second dose. Although any extrapolation into the future is uncertain, as yet there is no clear evidence to support recommendations of booster doses for those who have received two doses of an mRNA vaccine, other than vulnerable and immunosuppressed individuals who may require a third primary dose to achieve maximal protection [24].

## Declarations

### Public and Patient Involvement statement

This study was conducted under approvals from the Public Benefit and Privacy Panel for Health and Social Care which includes public and patient representatives.

### Ethics approval

This study was performed within Public Health Scotland as part of its statutory duty to monitor and investigate public health problems. Under the UK Policy Framework for Health and Social Care Research set out by the NHS Health Research Authority, this does not fall within the definition of research and ethical review was therefore not required. Individual consent is not required for Public Health Scotland staff to process personal data to perform specific tasks in the public interest that fall within its statutory role. The statutory basis for this is set out in Public Health Scotland’s privacy notice.

### Transparency declaration

PM as the manuscript’s guarantor affirms: that the manuscript is an honest, accurate, and transparent account of the study being reported; that no important aspects of the study have been omitted; and that any discrepancies from the study as originally planned and registered have been explained. This manuscript has been generated directly from the source data by a reproducible research pipeline.

### Author contributions

Conceptualization: all authors. Formal analysis: PM. Writing original draft: PM, HC. Review and editing: all authors.

### Data Availability

The component datasets used here are available via the Public Benefits and Privacy Panel for Health and Social Care at https://www.informationgovernance.scot.nhs.uk/pbpphsc/ for researchers who meet the criteria for access to confidential data. All source code used for derivation of variables, statistical analysis and generation of this manuscript is available on https://github.com/pmckeigue/covid-scotland_public.

## Acknowledgements

We thank Jen Bishop, Ciara Gribben, Bob Taylor and David Caldwell for undertaking the linkage analysis within Public Health Scotland,

## Supplementary Material

**Fig S1.**
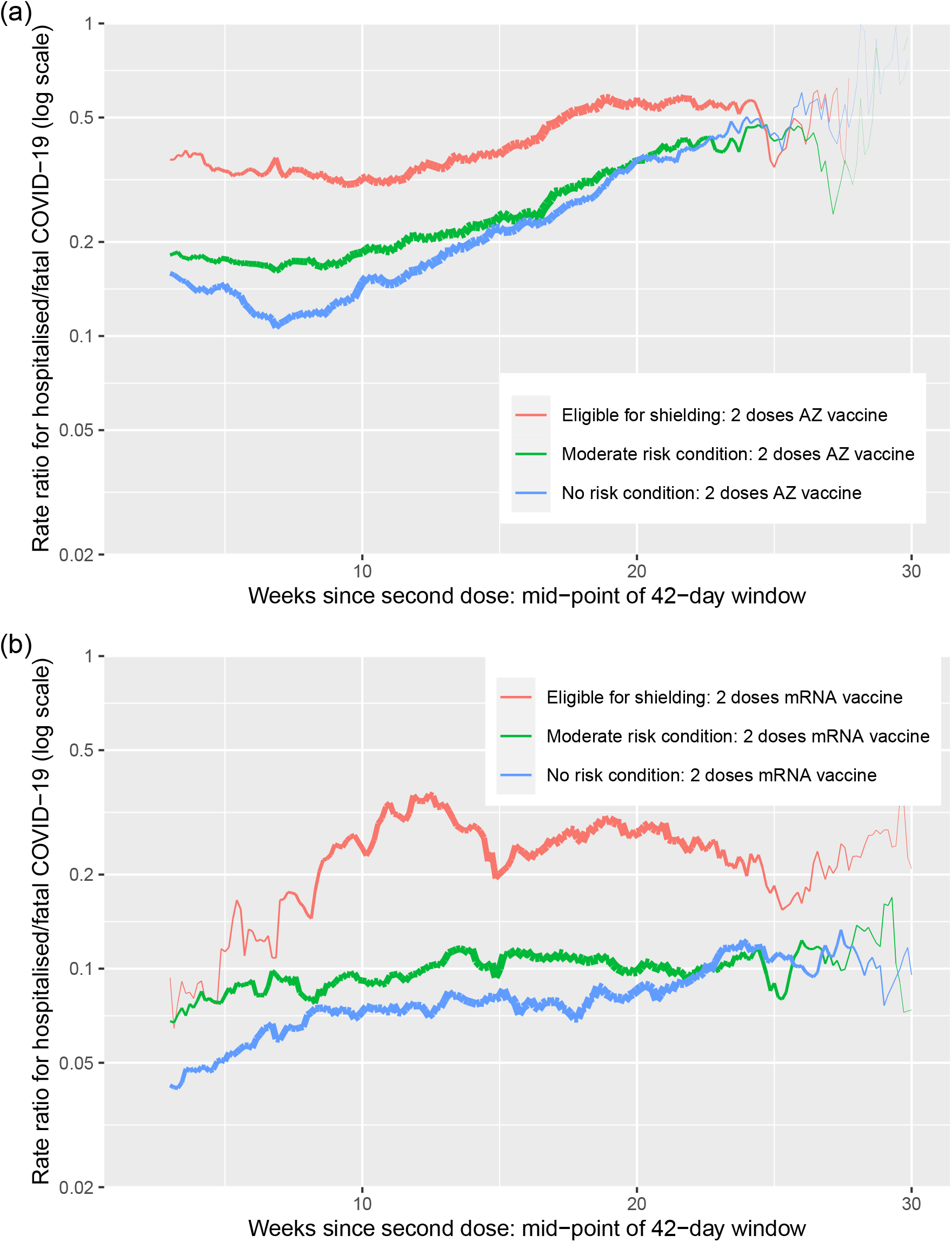
Relation of efficacy of two doses of vaccine against hospitalised or fatal COVID-19 to time since last dose by risk group: (a) AstraZeneca vaccine; (b) mRNA vaccines. Rate ratios in conditional logistic regression model, adjusted for covariates. For each effect, line thickness is proportional to precision of estimate

**Fig S2.**
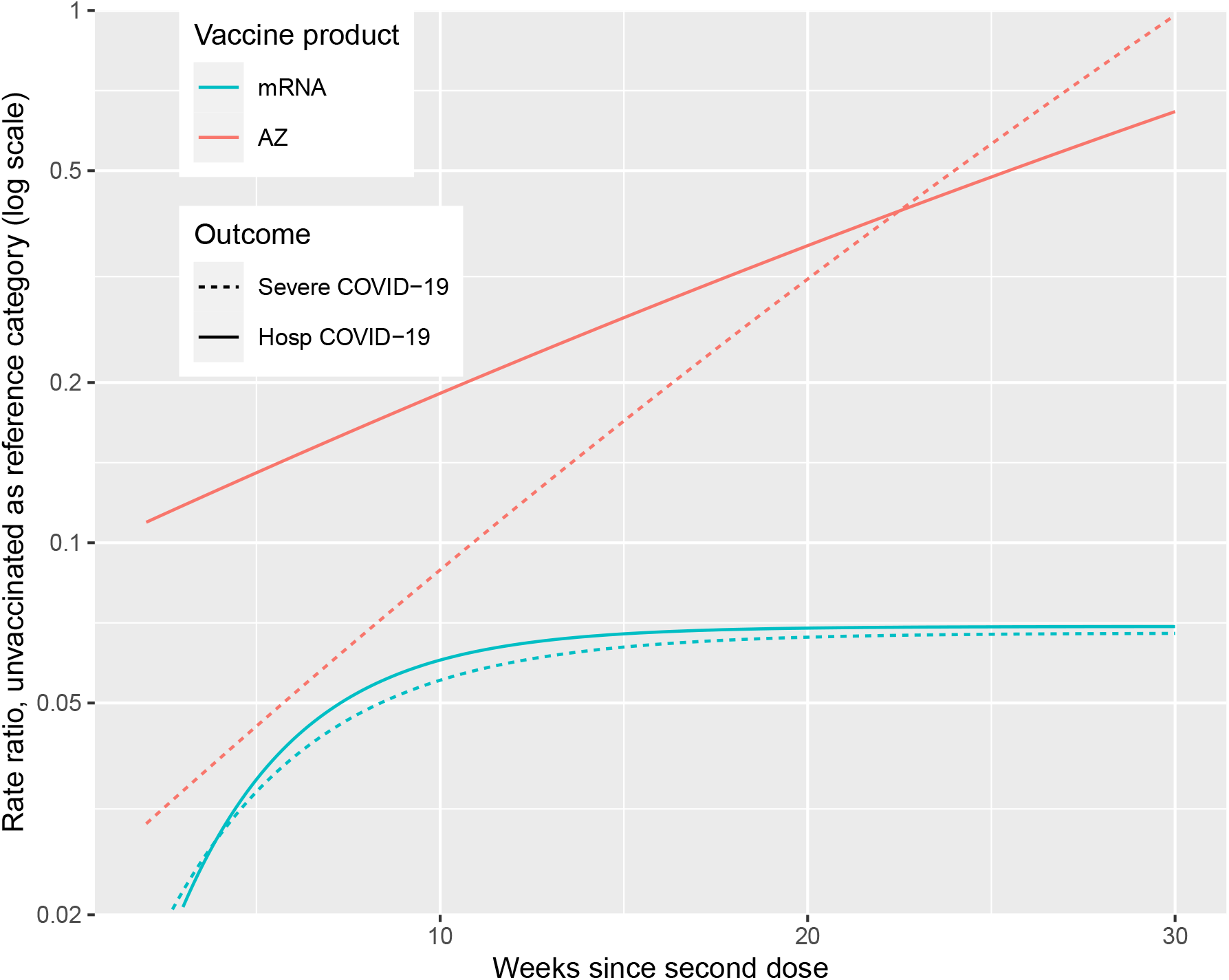
Best-fitting models of waning to constant efficacy for the narrow outcome of severe COVID-19 and the broader outcome of hospitalised or fatal COVID-19

**Table S1.** Comparison of models for waning of efficacy

| Waning model | Product | Outcome | Difference in log-likelihood | Decay half-life (days) |
| --- | --- | --- | --- | --- |
| To zero | AZ | Severe | 0.0 | 85 (95% CI 65 to 126) |
| To constant efficacy | AZ | Severe | 1.4 | >500 (95% CI 68 to Inf) |
| To zero | mRNA | Severe | 0.0 | >500 (95% CI 201 to Inf) |
| To constant efficacy | mRNA | Severe | 1.4 | 20 (95% CI 3 to Inf) |
| To zero | AZ | Hospitalised | 0.0 | 37 (95% CI 32 to 45) |
| To constant efficacy | AZ | Hospitalised | 1041.3 | >500 (95% CI 212 to Inf) |
| To zero | mRNA | Hospitalised | 0.0 | 116 (95% CI 67 to 397) |
| To constant efficacy | mRNA | Hospitalised | 224.4 | 16 (95% CI 4 to 101) |

Differences in log-likelihood > 11 natural log units are significant at $p < 0.001$
Conditional logistic regression models include care home residence, clinical risk category, number of non-cardiovascular drug classes and recent inpatient stay as covariates

**Table S2.** Rate ratios for severe COVID-19

|  | Controls (50086) | Cases (5644) | Univariate |  | Multivariable |  |
| --- | --- | --- | --- | --- | --- | --- |
|  |  |  | Rate ratio (95% CI) | *p*-value | Rate ratio (95% CI) | *p*-value |
| Care home | 2471 (5%) | 951 (17%) | 5.1 (4.6, 5.7) | $2 \times 10^{-210}$ | 5.1 (4.5, 5.8) | $2 \times 10^{-148}$ |
| Clinical risk category |  |  |  |  |  |  |
| No risk condition | 28661 (57%) | 1576 (28%) |  |  |  |  |
| Moderate risk condition | 17603 (35%) | 2891 (51%) | 3.31 (3.09, 3.55) | $7 \times 10^{-243}$ | 1.96 (1.80, 2.13) | $1 \times 10^{-57}$ |
| Eligible for shielding | 3822 (8%) | 1177 (21%) | 6.5 (5.9, 7.0) | $2 \times 10^{-371}$ | 2.96 (2.65, 3.31) | $1 \times 10^{-80}$ |
| Number of non-cardiovascular drug classes | 2 (1 to 5) | 6 (3 to 9) | 1.20 (1.20, 1.21) | $1 \times 10^{-538}$ | 1.12 (1.11, 1.13) | $6 \times 10^{-132}$ |
| Recent hospital stay | 1062 (2%) | 1479 (26%) | 17.1 (15.5, 18.8) | $2 \times 10^{-745}$ | 11.8 (10.6, 13.1) | $4 \times 10^{-478}$ |
| Vaccination status |  |  |  |  |  |  |
| Not vaccinated | 36874 (74%) | 4494 (80%) |  |  |  |  |
| 1 dose mRNA vaccine | 2277 (5%) | 247 (4%) | 0.68 (0.58, 0.81) | $6 \times 10^{-6}$ | 0.39 (0.33, 0.47) | $8 \times 10^{-24}$ |
| 1 dose AZ vaccine | 2693 (5%) | 260 (5%) | 0.39 (0.32, 0.47) | $2 \times 10^{-22}$ | 0.34 (0.27, 0.41) | $1 \times 10^{-24}$ |
| 2 doses mRNA vaccine | 2630 (5%) | 142 (3%) | 0.11 (0.09, 0.14) | $1 \times 10^{-71}$ | 0.07 (0.05, 0.09) | $3 \times 10^{-90}$ |
| 2 doses AZ vaccine | 5595 (11%) | 499 (9%) | 0.18 (0.15, 0.22) | $2 \times 10^{-75}$ | 0.13 (0.10, 0.15) | $5 \times 10^{-95}$ |

Presentation dates from 1 December 2020 to 19 August 2021.
Vaccination status defined as number of doses received at least 14 days before presentation date.
Controls matched for age, sex, and primary care practice on date of presentation of case.
Multivariable model includes all covariates in the table

**Table S3.** Rate ratios for hospitalised or fatal COVID-19

|  | Controls (202071) | Cases (21671) | Univariate |  | Multivariable |  |
| --- | --- | --- | --- | --- | --- | --- |
|  |  |  | Rate ratio (95% CI) | *p*-value | Rate ratio (95% CI) | *p*-value |
| Care home | 5790 (3%) | 2053 (9%) | 4.27 (4.00, 4.56) | $2 \times 10^{-410}$ | 3.32 (3.06, 3.61) | $2 \times 10^{-184}$ |
| Clinical risk category |  |  |  |  |  |  |
| No risk condition | 133911 (66%) | 8802 (41%) |  |  |  |  |
| Moderate risk condition | 55806 (28%) | 9214 (43%) | 2.98 (2.88, 3.09) | $2 \times 10^{-817}$ | 1.82 (1.75, 1.90) | $4 \times 10^{-175}$ |
| Eligible for shielding | 12354 (6%) | 3655 (17%) | 5.6 (5.3, 5.8) | $2 \times 10^{-1125}$ | 2.61 (2.46, 2.78) | $5 \times 10^{-219}$ |
| Number of non-cardiovascular drug classes | 2 (0 to 4) | 4 (2 to 8) | 1.19 (1.19, 1.20) | $2 \times 10^{-1762}$ | 1.12 (1.12, 1.13) | $3 \times 10^{-490}$ |
| Recent hospital stay | 3304 (2%) | 5333 (25%) | 21.3 (20.3, 22.5) | $3 \times 10^{-2850}$ | 14.7 (13.9, 15.5) | $7 \times 10^{-1922}$ |
| Vaccination status |  |  |  |  |  |  |
| Not vaccinated | 128417 (64%) | 15369 (71%) |  |  |  |  |
| 1 dose mRNA vaccine | 11812 (6%) | 788 (4%) | 0.37 (0.34, 0.41) | $3 \times 10^{-110}$ | 0.30 (0.27, 0.33) | $2 \times 10^{-143}$ |
| 1 dose AZ vaccine | 12562 (6%) | 1440 (7%) | 0.59 (0.55, 0.65) | $4 \times 10^{-34}$ | 0.50 (0.46, 0.55) | $2 \times 10^{-51}$ |
| 2 doses mRNA vaccine | 16983 (8%) | 774 (4%) | 0.13 (0.12, 0.15) | $1 \times 10^{-348}$ | 0.10 (0.09, 0.11) | $10 \times 10^{-401}$ |
| 2 doses AZ vaccine | 32213 (16%) | 3294 (15%) | 0.33 (0.31, 0.36) | $2 \times 10^{-188}$ | 0.22 (0.21, 0.24) | $4 \times 10^{-291}$ |

Presentation dates from 1 December 2020 to 19 August 2021.
Vaccination status defined as number of doses received at least 14 days before presentation date.
Controls matched for age, sex, and primary care practice on date of presentation of case.
Multivariable model includes all covariates in the table

## References

1. Nanduri S. Effectiveness of Pfizer-BioNTech and Moderna Vaccines in Preventing SARS-CoV-2 Infection Among Nursing Home Residents Before and During Widespread Circulation of the SARS-CoV-2 B.1.617.2 (Delta) Variant, March 1August 1, 2021. MMWR Morbidity and Mortality Weekly Report. 2021;70. doi:10.15585/mmwr.mm7034e3

2. Lopez Bernal J, Andrews N, Gower C, Gallagher E, Simmons R, Thelwall S, et al. Effectiveness of Covid-19 Vaccines against the B.1.617.2 (Delta) Variant. New England Journal of Medicine. 2021;385: 585–594. doi:10.1056/NEJMoa2108891

3. Pouwels KB, Pritchard E, Matthews PC, Stoesser N, Eyre DW, Vihta K-D, et al. Impact of Delta on viral burden and vaccine effectiveness against new SARS-CoV-2 infections in the UK. Cold Spring Harbor Laboratory Press; 2021. p. 2021.08.18.21262237. doi:10.1101/2021.08.18.21262237

4. Tenforde MW. Sustained Effectiveness of Pfizer-BioNTech and Moderna Vaccines Against COVID-19 Associated Hospitalizations Among Adults, March – July 2021. MMWR Morbidity and Mortality Weekly Report. 2021;70. doi:10.15585/mmwr.mm7034e2

5. Rosenberg ES. New COVID-19 Cases and Hospitalizations Among Adults, by Vaccination Status, May 3 – July 25, 2021. MMWR Morbidity and Mortality Weekly Report. 2021;70. doi:10.15585/mmwr.mm7034e1

6. Puranik A, Lenehan PJ, Silvert E, Niesen MJM, Corchado-Garcia J, O’Horo JC, et al. Comparison of two highly-effective mRNA vaccines for COVID-19 during periods of Alpha and Delta variant prevalence. Cold Spring Harbor Laboratory Press; 2021. p. 2021.08.06.21261707. doi:10.1101/2021.08.06.21261707

7. Mizrahi B, Lotan R, Kalkstein N, Peretz A, Perez G, Ben-Tov A, et al. Correlation of SARS-CoV-2 Breakthrough Infections to Time-from-vaccine; Preliminary Study. Cold Spring Harbor Laboratory Press; 2021. p. 2021.07.29.21261317. doi:10.1101/2021.07.29.21261317

8. Public Health England. Duration of protection of COVID-19 vaccines against clinical disease, 9 September 2021. Public Health England; 2021 Sep.

9. Office of the Commissioner. Joint Statement from HHS Public Health and Medical Experts on COVID-19 Booster Shots. FDA. https://www.fda.gov/news-events/press-announcements/joint-statement-hhs-public-health-and-medical-experts-covid-19-booster-shots; FDA; 2021.

10. Public Health England. JCVI issues updated advice on COVID-19 booster vaccination. http://GOV.UK. https://www.gov.uk/government/news/jcvi-issues-updated-advice-on-covid-19-booster-vaccination; 2021.

11. McKeigue PM, Weir A, Bishop J, McGurnaghan SJ, Kennedy S, McAllister D, et al. Rapid Epidemiological Analysis of Comorbidities and Treatments as risk factors for COVID-19 in Scotland (REACT-SCOT): A population-based case-control study. PLOS Medicine. 2020;17: e1003374. doi:10.1371/journal.pmed.1003374

12. McKeigue PM, Kennedy S, Weir A, Bishop J, McGurnaghan SJ, McAllister D, et al. Relation of severe COVID-19 to polypharmacy and prescribing of psychotropic drugs: The REACT-SCOT case-control study. BMC medicine. 2021;19: 51. doi:10.1186/s12916-021-01907-8

13. McKeigue PM, McAllister DA, Caldwell D, Gribben C, Bishop J, McGurnaghan S, et al. Relation of severe COVID-19 in Scotland to transmission-related factors and risk conditions eligible for shielding support: REACT-SCOT case-control study. BMC medicine. 2021;19: 149. doi:10.1186/s12916-021-02021-5

14. Public Health Scotland. COVID-19 vaccination in Scotland: Daily update. COVID-19 Daily Dashboard. https://public.tableau.com/views/COVID-19DailyDashboard_15960160643010/Overview?/; 2021.

15. Sheikh A, McMenamin J, Taylor B, Robertson C. SARS-CoV-2 Delta VOC in Scotland: Demographics, risk of hospital admission, and vaccine effectiveness. The Lancet. 2021;397: 2461–2462. doi:10.1016/S0140-6736(21)01358-1

16. Breslow NE, Day NE, Halvorsen KT, Prentice RL, Sabai C. Estimation of multiple relative risk functions in matched case-control studies. American Journal of Epidemiology. 1978;108: 299–307. doi:10.1093/oxfordjournals.aje.a112623

17. Greenland S, Thomas DC. On the need for the rare disease assumption in case-control studies. American Journal of Epidemiology. 1982;116: 547–553. doi:10.1093/oxfordjournals.aje.a113439

18. Public Health England. SARS-CoV-2 variants of concern and variants under investigation. 2021; 69.

19. Doria-Rose N, Suthar MS, Makowski M, O’Connell S, McDermott AB, Flach B, et al. Antibody Persistence through 6 Months after the Second Dose of mRNA-1273 Vaccine for Covid-19. New England Journal of Medicine. 2021;384: 2259–2261. doi:10.1056/NEJMc2103916

20. Andrews N, Tessier E, Stowe J, Gower C, Kirsebom F, Simmons R, et al. Vaccine effectiveness and duration of protection of Comirnaty, Vaxzevria and Spikevax against mild and severe COVID-19 in the UK. Public Health England; 2021 Sep. doi:10.1101/2021.09.15.21263583

21. Tartof SY, Slezak JM, Fischer H, Hong V, Ackerson BK, Ranasinghe ON, et al. Effectiveness of mRNA BNT162b2 COVID-19 vaccine up to 6 months in a large integrated health system in the USA: A retrospective cohort study. The Lancet. 2021;0. doi:10.1016/S0140-6736(21)02183-8

22. Krause PR, Fleming TR, Peto R, Longini IM, Figueroa JP, Sterne JAC, et al. Considerations in boosting COVID-19 vaccine immune responses. The Lancet. 2021;0. doi:10.1016/S0140-6736(21)02046-8

23. Barclay W, Kellam P, Moss P, Lopez-Bernal J, Ramsey M, Zambon M. How long will vaccines continue to protect against COVID-19?, 30 July 2021. Scientific Advisory Group for Emergencies; 2021 Jul.

24. Joint Committee on Vaccination and Immunisation. Joint Committee on Vaccination and Immunisation (JCVI) advice on third primary dose vaccination. https://www.gov.uk/government/publications/third-primary-covid-19-vaccine-dose-for-people-who-are-immunosuppressed-jcvi-advice/joint-committee-on-vaccination-and-immunisation-jcvi-advice-on-third-primary-dose-vaccination: Joint Committee on Vaccination and Immunisation; 2021 Sep.

